# Sensory symptoms across the lifespan in people with cerebral palsy

**DOI:** 10.1101/2023.07.21.23292955

**Authors:** Ariel M Lyons-Warren, Danielle Guez-Barber, Sruthi P. Thomas, Evelyne K Tantry, Aditya Mahat, Bhooma Aravamuthan, Cerebral Palsy Research Network

**Author notes:** **Corresponding Author:** Bhooma R. Aravamuthan, Division of Pediatric Neurology, Department of Neurology Washington University School of Medicine, 660 South Euclid Avenue Campus Box 8111, St. Louis, MO, 63110-1093, USA.

## Abstract

**AIM:** To estimate prevalence of sensory symptoms in people with cerebral palsy (CP) across the lifespan.

**METHOD:** In this cross-sectional study, the self-reported Sensory Processing Scale Inventory (SPS-I) was administered via REDCap between 2/1/22 and 8/15/22 to people with CP or their caregivers enrolled in the online MyCP Community Registry. We determined the association between SPS-I scores and age (Pearson correlation) and functional status as assessed using five validated functional classification systems for CP (ANOVA). We hypothesized that sensory symptoms would differ between younger and older individuals with CP.

**RESULTS:** Of 155 responses (28% response rate, age 1-76 years, 34% male), 97% reported at least one bothersome sensory symptom. Total sensory symptoms decreased with age (R^2^=0.12, p<0.0001), driven by decreases in hyposensitivity symptoms (R^2^=0.32, p<0.0001), primarily tactile hyposensitivity (R^2^=0.29, p<0.0001). Sensory symptoms increased with greater functional impairment across all functional domains (ANOVA, p<0.0001). However, the age-specific decrease in hypo-sensitivities was most pronounced in people with the greatest gross motor functional impairment (R^2^=0.70, p=0.0004).

**INTERPRETATION:** Our findings suggest that hypo-sensitivity, primarily tactile sensitivity, decreases with age in people with CP. Future work should assess whether decreased hyposensitivity contributes to other age-related changes in CP like increased pain.

**What this paper adds:**

- Individuals with CP exhibit both hyper- and hypo-sensitivity across all sensory modalities
- Hyposensitivity decreases with age in people with CP, independent of GMFCS level
- Less hyposensitivity correlates with greater functional ability in people with CP
- Of all sensory modalities, tactile hyposensitivity correlates most strongly with age.

---

Cerebral palsy (CP) is the most common cause of disability in childhood affecting up to 1 in 250 children in the United States (https://www.cdc.gov/ncbddd/cp/data.html). Children with CP develop motor difficulties secondary to non-progressive early changes in the brain, either through genetic factors or injury in the prenatal, perinatal, or early postnatal period. Separate from motor disabilities, individuals with CP have highly heterogeneous involvement of the sensory system^1^. Sensory symptoms experienced by individuals with CP include both hyper (more) or hypo (less) sensitivity to touch, sight, sound, taste, smell or proprioceptive awareness. These symptoms are typically assessed by self or caregiver report^2^. For example, covering one’s ears in response to sound suggests auditory hypersensitivity. Sensory symptoms can also be directly measured by quantitative sensory testing in the clinic^3–4^.

Sensory symptoms in children with CP are common. Tactile deficits specifically have been well described^5^. Using the Sensory Profile questionnaire, children with CP (N=43) exhibited differences in more than half of assessed categories including visual, vestibular, and multisensorial processing compared to typically developing controls (N=59)^6^. Sensory symptoms are also associated with functional ability in self-care^7^ and mobility^8^. Multiple small studies (N 9 to 30) using clinical measures of sensory symptoms in children with CP found deficits in proprioception, stereognosis, tactile perception, and tactile discrimination^5,9,10^. Similarly, it is well established that individuals with CP have increased pain and pain perception compared to the typically developing population^11,12^. Despite increasing recognition of sensory symptoms in CP, neither sensory symptoms across the lifespan nor discriminating hyper and hypo sensitivity in CP have been well characterized.

The initial brain injury or neurodevelopmental disruption that causes CP does not change with age, but it has been well established that the phenotypic manifestations of CP evolve over the lifespan. Gross motor function declines with age and this decline can even begin mid-childhood for those children who are more functionally impaired^13^. Concomitantly, pain is highly prevalent in the adult population with CP and seems to increase with age and level of motor impairment^14^. A significant percentage of children with CP have dysphagia and at least one study indicates that it can increase in severity with age^15^. Given all age-related changes in the presentation of CP, we hypothesized there are also differences in sensory symptoms across the lifespan.

Thus, sensory symptoms are an important yet understudied component of CP with significant gaps in our understanding across sensory modalities and the lifespan. To address these gaps, we conducted a large-scale characterization of sensory differences in individuals with CP across the lifespan using the Sensory Processing Scale Inventory (copyright Miller and Schoen 2004 and 2005, SPS-I)^16^, a comprehensive sensory assessment questionnaire. We hypothesized that sensory symptoms would differ between younger and older individuals with CP. Determining these potentially age-related differences could lay the groundwork for future studies determining how sensory symptoms may underlie the functional changes seen in people with CP as they age.

## Methods

This cross-sectional survey study received a human subject research exemption from the Washington University Institutional Review Board (IRB Identification Number: 201908233).

Caregivers and people with CP were invited to participate based on their enrollment in MyCP via the Cerebral Palsy Research Network (CPRN). The CPRN community registry gathers data directly from people with CP and their caregivers via REDCap-based surveys distributed to the community through MyCP.org^17^. People with CP and their caregivers sign-up to participate through the MyCP.org portal and are presented with surveys for which they meet the designated inclusion criteria. All MyCP participants were presented with this survey. The registrant’s CP diagnosis is ascertained by a standardized medical history intake form. The survey for this study used the SPS-I which has both adult^18^ and pediatric^19^ versions to assess sensory symptoms in people with CP either by self-report or proxy report by a primary caregiver. The survey also asked for information about each participant’s functional status as determined using the five validated functional classification systems for CP: Gross Motor Function Classification System (GMFCS)^20^, Manual Ability Classification System (MACS)^21^, Eating and Drinking Ability Classification System (EDACS)^22^, Communication Function Classification System (CFCS)^23^, and Visual Function Classification System (VFCS)^24^. The survey was integrated into the MyCP portal and available to MyCP registrants between 2/1/22 and 8/15/22. Incomplete surveys were excluded from the analysis.

The adult SPS-I, validated for ages 15 to 76, is comprised of 107 questions divided into six hypersensitivity sections: tactile, vision, olfactory, gustatory, auditory, and proprioception, and five hyposensitivity sections: tactile, olfactory, vision, auditory and proprioception. The pediatric SPS-I, validated for ages 1 to 14, is comprised of 68 questions divided into the same six hypersensitivity sections, but only three hyposensitivity sections: tactile, vision, and auditory. For each question, the respondent is asked to identify yes or no if an object or sensory experience is bothersome. During initial validation, typically developing children answered “yes” to less than 1 question on the total SPS-I compared to 5-8 “yes” responses for the clinical group.^16^ In this study, the number of yes responses within each section were summed and divided by the number of questions within that section to achieve a score between 0 (no sensory symptoms) to 1 (endorsed all queried sensory symptoms) to facilitate comparison across sensory modalities. Scores were calculated across all sensory modalities for hyposensitivities, hypersensitivies, and across both hypo- and hypersensitivies (called the total SPS-I score). Separate scores were also calculated for hyposensitivities, hypersensitivies, and across both hypo- and hypersensitivies for each sensory modality. All scores are reported on a 0-1 scale.

Of note, the pediatric SPS-I does not include questions on olfactory or proprioceptive hyposensitivity. Further, several adult hypersensitivity sections contained questions not applicable to pediatric patients. One question from the pediatric SPS-I “The person appears to be in their own world (tuned out)” could not be categorized by sensory modality. Thus, in order to directly compare pediatric and adult scores, our analysis included only a subset of questions. Specifically, the non-categorizable pediatric question was excluded and adult sensory questions that assessed similar concerns were consolidated so that the number of questions in the adult and pediatric hypersensitivity sections were comparable (Supplementary Table 1, survey instrument in Supplementary Methods). Participants who answered yes to any of the consolidated questions were marked as “yes” for the combined question. Participants answered all questions from the original SPS-I; question adjustment was made during analysis.

### Statistical Analyses

All statistical analysis were done in GraphPad Prism (version 8, GraphPad Software). Age (as a continuous variable) and SPS-I scores were compared between different functional classification system levels using ANOVAs with post-hoc Tukey tests (e.g. the difference in age between respondents at GMFCS levels I, II, III, IV, and V were compared using an ANOVA). We assessed for linear correlations between age and SPS-I scores using Pearson correlations. We assessed whether age or functional classification system level across the five assessed functional domains could predict the SPS-I score in aggregate across all sensory modalities using multiple linear regression, with separate assessment of the overall hyposensitivity score, overall hyposensitivity score, and overall SPS-I score including both hypo- and hypersensitivities. We assumed variables were independent (lack of multicollinearity) for variance inflation factors (VIFs) less than 5^25^. VIFs were calculated using collinearity diagnostics in the linear regression statistics package in GraphPad Prism. The significance level for all tests was set a priori to p<0.05.

## Results

### Participants

Out of 554 participants in the MyCP registry, we received responses from 180 individuals including 139 participants ages 15-76 who took the “adult” survey of whom 9 were age 15-18 and 41 participants ages 1-14 who took the “pediatric” survey. However, 25 participants (12 who took the adult instrument and 13 who completed the pediatric instrument) were excluded for incomplete responses, yielding 155 responses overall. 64% (98) of participants were female, 34% (52) were male and 2% (4) identified as non-binary or transgender. Age of included participants spanned 1 to 76 years of age (mean and standard deviation 38.28 ± 20.71) with variable levels of function (Table 1). As expected, self-respondents were older (median age 48, range 16-76, N=112) than the individuals with CP being described by caregivers (median age 11, range 2-57. N=43).

**Table 1:**
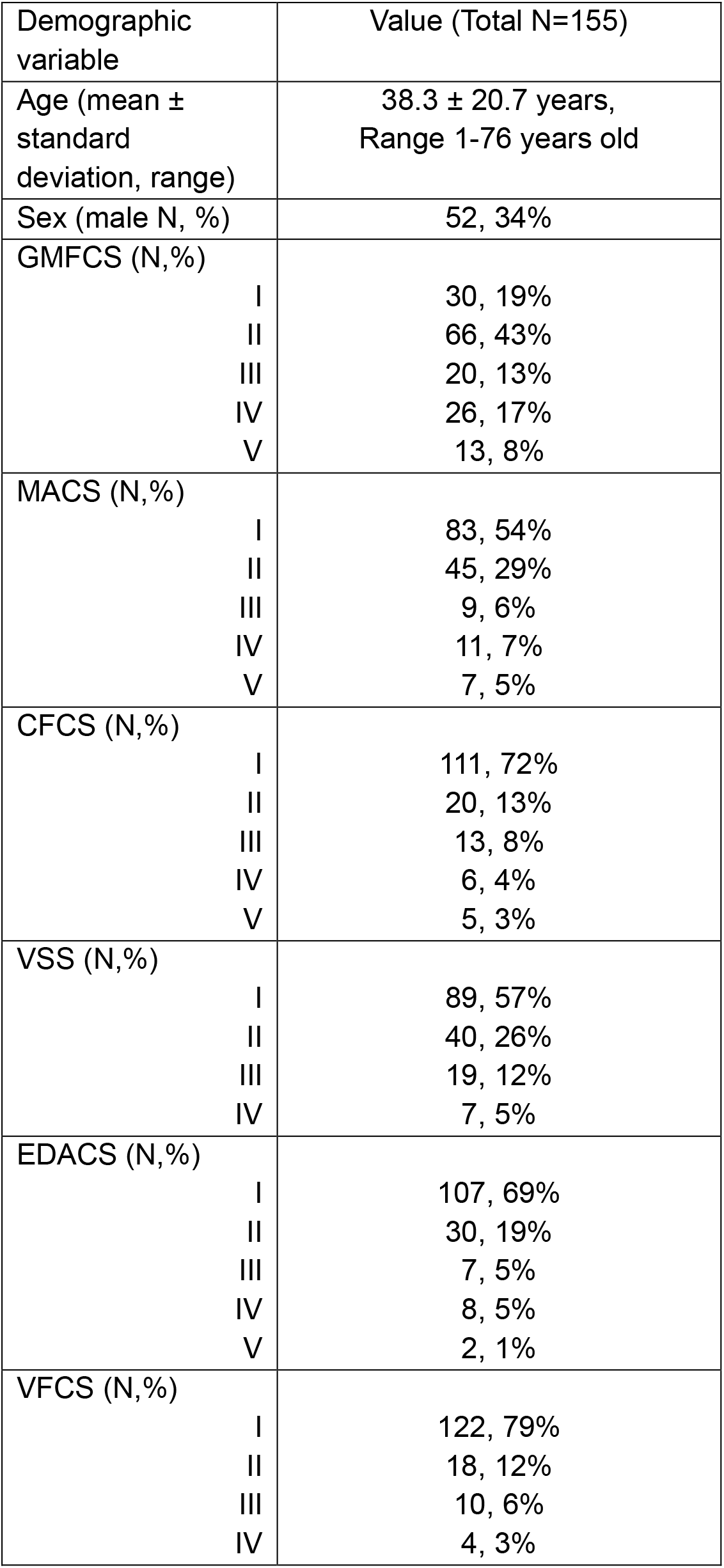
Distribution of Functional Scores; Number (N) of participants at each functional level for GMFCS, MACS, CCS, VSS, EDACS and VFCS including the mean score for each functional domain.

| Demographic variable | Value (Total N=155) |
| --- | --- |
| Age (mean ± standard deviation, range) | 38.3 ± 20.7 years, Range 1-76 years old |
| Sex (male N, %) | 52, 34% |
| GMFCS (N,%) |  |
| I | 30, 19% |
| II | 66, 43% |
| III | 20, 13% |
| IV | 26, 17% |
| V | 13, 8% |
| MACS (N,%) |  |
| I | 83, 54% |
| II | 45, 29% |
| III | 9, 6% |
| IV | 11, 7% |
| V | 7, 5% |
| CFCS (N,%) |  |
| I | 111, 72% |
| II | 20, 13% |
| III | 13, 8% |
| IV | 6, 4% |
| V | 5, 3% |
| VSS (N,%) |  |
| I | 89, 57% |
| II | 40, 26% |
| III | 19, 12% |
| IV | 7, 5% |
| EDACS (N,%) |  |
| I | 107, 69% |
| II | 30, 19% |
| III | 7, 5% |
| IV | 8, 5% |
| V | 2, 1% |
| VFCS (N,%) |  |
| I | 122, 79% |
| II | 18, 12% |
| III | 10, 6% |
| IV | 4, 3% |

MyCP registrants had significantly different functional statuses based on age, noting that younger people in the registry tended to have greater functional limitations (i.e. higher functional classification system levels) than older people in the registry (p<0.0001, ANOVA) (Figure 1). For GMFCS level, respondents at GMFCS level V were significantly younger than any of the respondents at GMFCS levels I-IV, without any significant difference in age distributions between GMFCS levels I-IV. For MACS, EDACS, CFCS, and VFCS, respondents at level I were significantly older than respondents at levels II-V, without any significant difference in age distributions between levels II-V. Given this, the assessment of differences in SPS-I relative to age was done taking these group differences in functional status into account. That is, the SPS-I scores were compared to the age of the respondents for those at GMFCS levels I-IV separately from respondents at GMFCS level V as a secondary analysis. Similarly, a secondary analysis comparing SPS-I scores to age for respondents at MACS, EDACS, CFCS, and VFCS level I separately from respondents at levels II-V was done.

**Figure 1:**
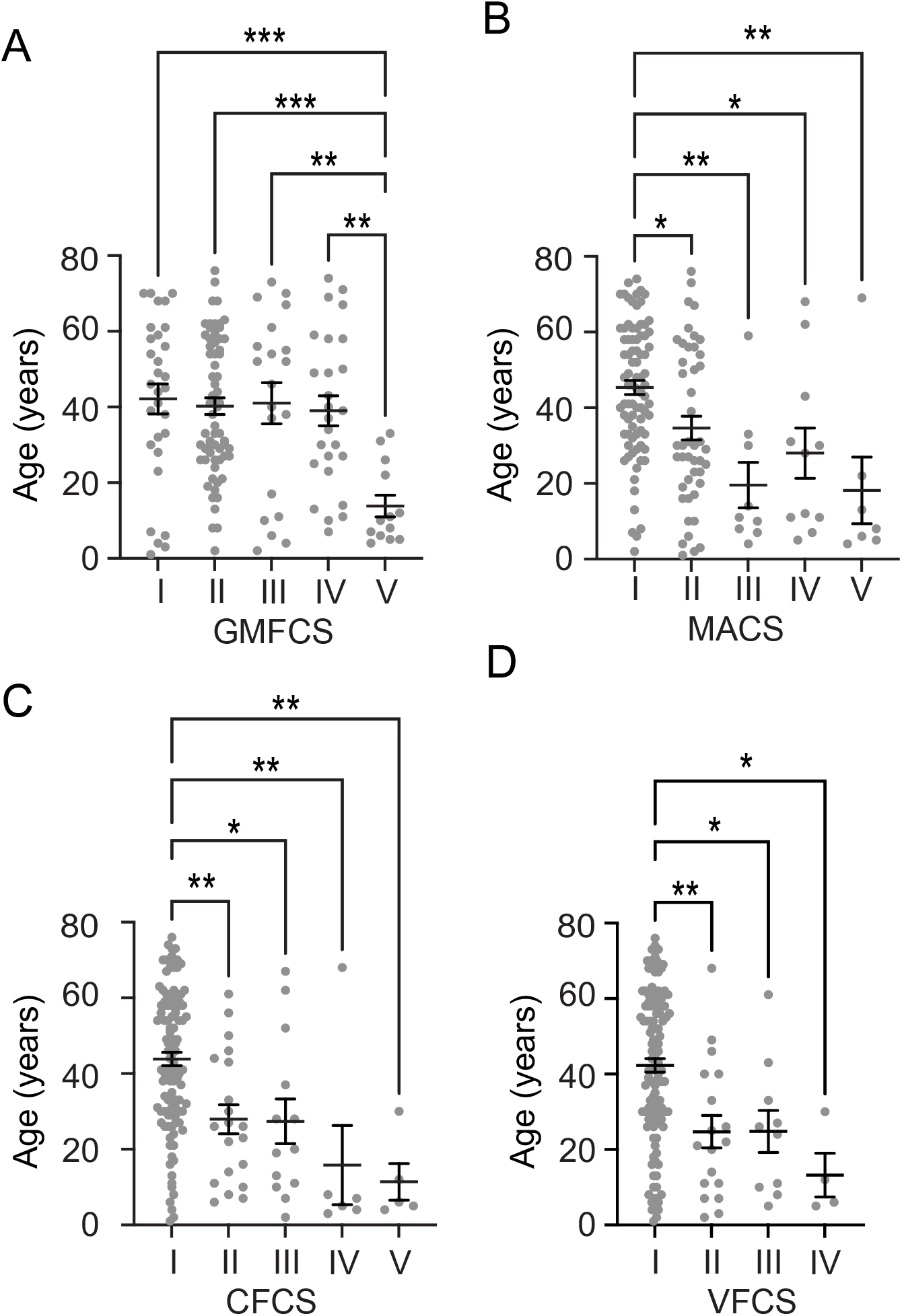
Functional Status varied by age. Participant age plotted by GMFCS (A), MACS (B), CFCS (C) and VFCS (D) shows a significantly lower mean age associated with higher numerical levels (greater functional impairment). Comparisons are all by ANOVA with significant differences between groups shown as *p<0.05, **p<0.01, ***p<0.005. Of note, ages were compared across all levels within each functional classification system, but only significant differences in age between levels are shown.

### Individuals with CP exhibit sensory symptoms in all modalities

Consistent with prior, smaller studies^6–8^, we observed that almost all our participants reported sensory symptoms above the typical range^16^ (Figure 2A). Interestingly, we also saw a wide distribution of scores for touch (Figure 2B), vision (Figure 2C), auditory (Figure 2D) and proprioception (Figure 2E). While 5 (3%) subjects reported no sensory symptoms in any area, almost all participants reported at least one sensory symptom and 30 (19%) reported at least one sensory symptom in each sensory modality.

**Figure 2:**
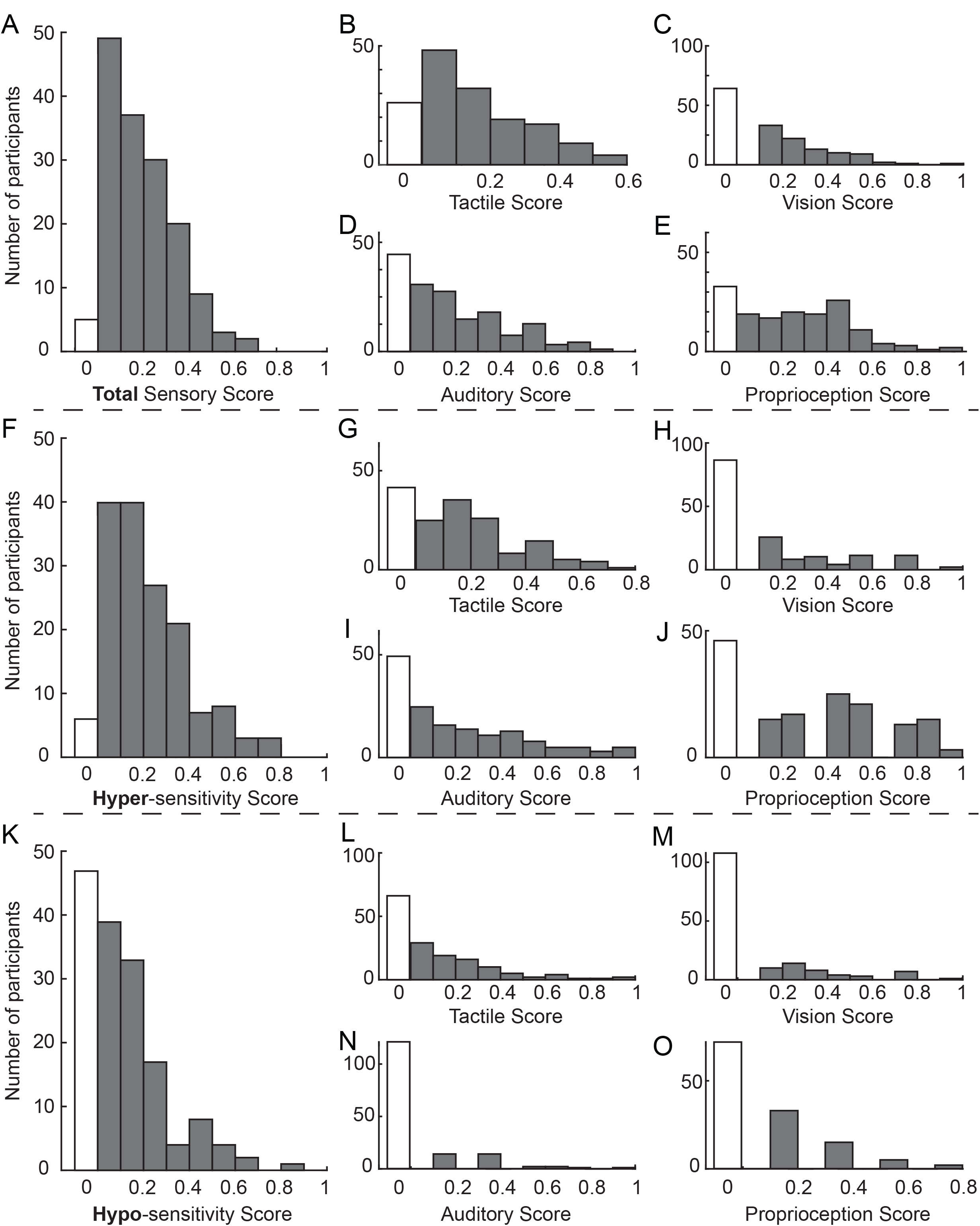
Distribution of Sensory Symptoms by type of sensitivity and sensory modality Histograms showing the number of participants with each score range for total sensory score (A), total tactile score (B), total vision score (C), total auditory score (D), total proprioception score (E), total hyper-sensitivity score (F), hyper-sensitivity tactile score (G), hyper-sensitivity vision score (H), hyper-sensitivity auditory score (I), hyper-sensitivity proprioception score (J), total hypo-sensitivity score (K), hypo-sensitivity tactile score (L), hypo-sensitivity vision score (M), hypo-sensitivity auditory score (N), and hypo-sensitivity proprioception score (O). White bars indicate number of individuals who answered no to all questions in that category for a score of 0.

### Individuals with CP exhibit both hyper and hypo sensitivity symptoms

Similar to both typically developing children and other clinical populations^16^, hypersensitivity symptoms were more common than hyposensitivity symptoms in people with CP. 149/155 (96%) of participants noted at least one hyper-sensitivity symptom, with a comparable frequency distribution for hypersensitivity SPS-I scores as was seen for total SPS-I scores (Figure 2F), including within each sensory modality (Figure 2G-J). Fewer respondents, though still a majority, reported at least one hypo-sensitivity symptom (108/155, 70%) (Figure 2K-O).

Hyper- and hypo-sensitivity symptoms co-existed in the same person with CP. The majority (107/155, 69%) of participants reported at least one sensory symptom for both hyper and hypo sensitivity. Of the 48 participants who did not report both hyper and hypo sensitivity symptoms, 5 reported no sensory symptoms, 42 reported only hyper-sensitivity symptoms, and 1 reported only hypo-sensitivity symptoms.

Thus, individuals with CP exhibit a range of sensory symptoms across all sensory modalities and are impacted by both hypo- and hyper-sensitivity.

### A decrease in tactile hypo-sensitivity specifically is the strongest driver of age-related differences in sensory symptoms

Next, we were interested in how sensory symptoms differ by age. We looked at total sensory score by age (Figure 3A) and found a trend towards less sensory symptoms in older participants (R^2^ = 0.12, p<0.0001). Interestingly, this trend was driven by changes in hypo-sensitivity (R^2^ = 0.32, p < 0.0001, Figure 3B) more so than hyper-sensitivity (R^2^ = 0.04, p = 0.04, Figure 3C). Given that there was a difference in age between the people with CP described by self-respondents and caregiver respondents and noting that caregiver respondents were able to respond for people with CP across the pediatric to adult age spectrum while self-respondents were not, we additionally assessed the relationship between age and sensory scores within only caregiver respondents. Similar relationships between age and sensory score were reflected within the caregiver respondents alone: caregivers reported decreased sensory symptoms overall in older people with CP (R^2^ = 0.31, p=0.04), which tended to be driven by reports of decreased hyposensitivity (R^2^ = 0.30, p=0.05) more so than decreased hypersensitivity (R^2^ = 0.10, p=0.50).

**Figure 3:**
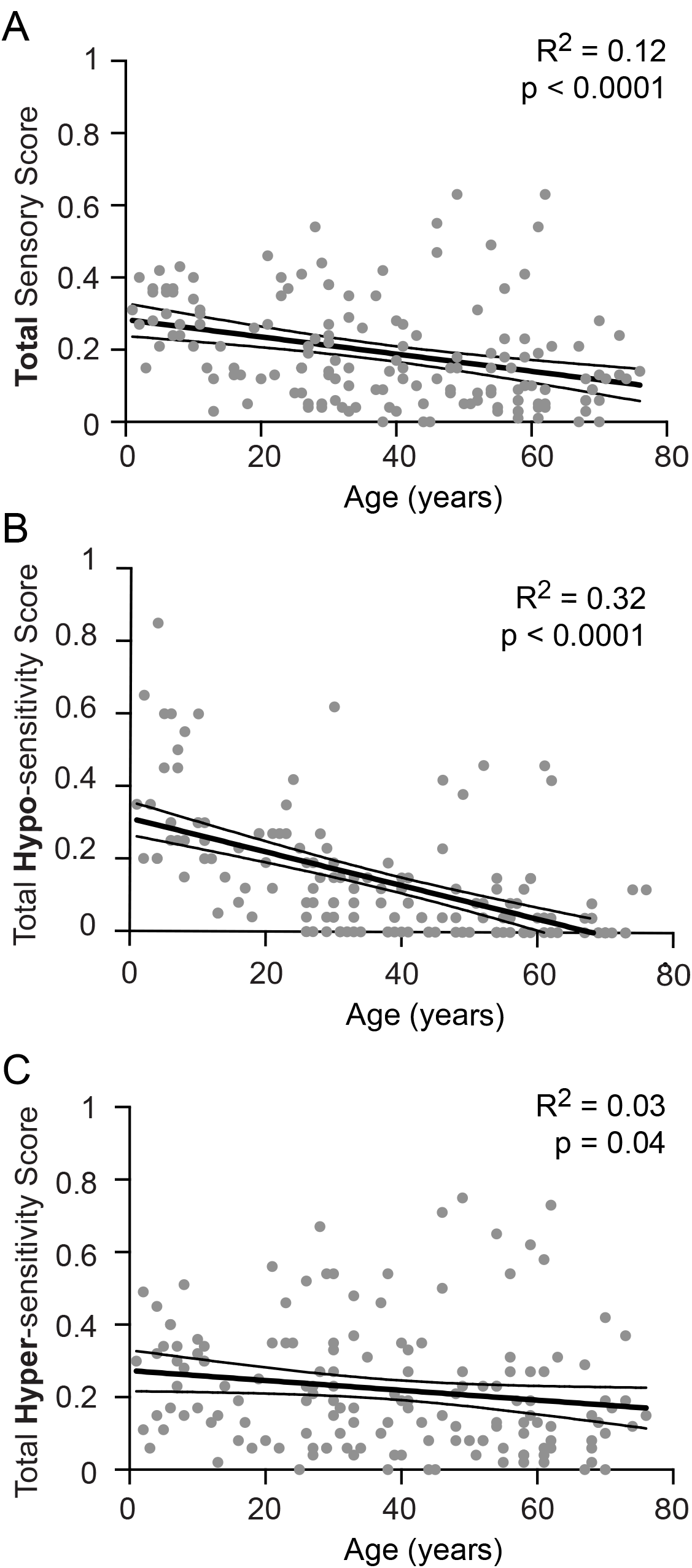
Sensory symptoms decrease with age. Total sensory (A), Hypo-sensitivity (B) and Hyper-sensitivity (C) scores decreased with age. Pearson correlation, *p<0.05.

Given that sensory changes have previously been correlated with functional ability in children^7–8^, we next evaluated sensory scores based on functional category. Indeed, hypo-sensitivity scores in our cohort were significantly higher in individuals with GMFCS V compared to all other functional categories (Figure 4A). Similarly, hypo-sensitivity scores were significantly lower in individuals with MACS I (Figure 4B), CFCS I (Figure 4C), and VFCS I (Figure 4D). Thus, the degree of sensory symptoms is positively associated with the degree of functional impairment for multiple areas of functional ability.

**Figure 4:**
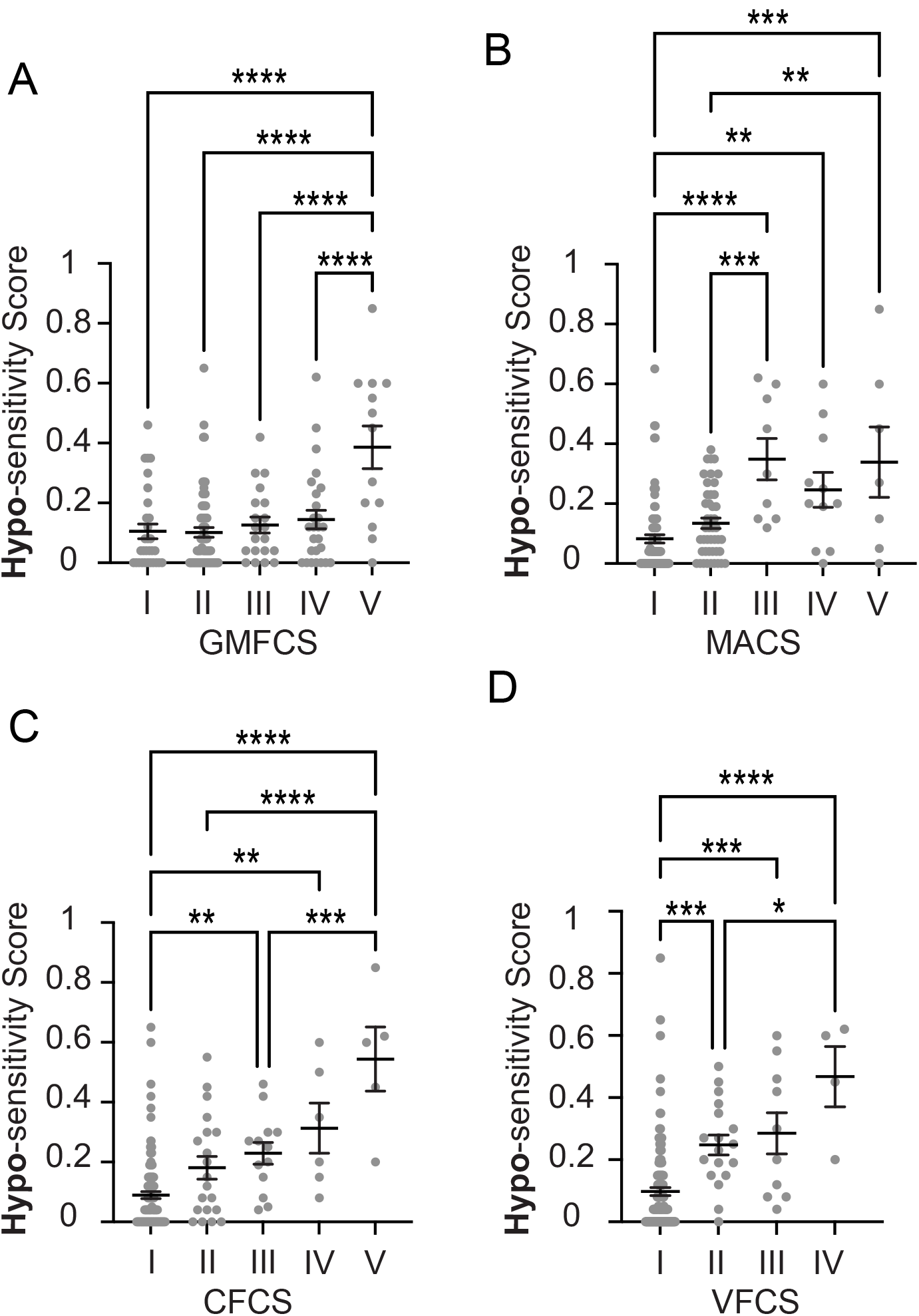
Hypo-sensitivity scores correlated with functional ability. Hypo-sensitivity scores were significantly different for individuals with GMFCS level V (A), MACS level V (B), CFCS level V (C) and VFCS level IV (D). Comparisons are all by ANOVA with significant differences between groups shown as *p<0.05, **p<0.01, ***p<0.005. Of note, hypo-sensitivity scores were compared across all levels within each functional classification system, but only significant differences in scores between levels are shown.

As noted above, we split participants based on functional level and reassessed the correlation between sensory score and age. Notably, the trend towards less sensory symptoms in older participants held for individuals at GMFCS I-IV (Figure 5A, R^2^ = 0.10, p<0.0001) and GMFCS V (Figure 5B, R^2^ = 0.61, p=0.002), with R^2^ values suggesting that the correlation between age and sensory symptoms were most pronounced in people with the greatest functional limitations. Again, this trend was strongest for hypo-sensitivity (Figure 5C,D) but still present for hyper-sensitivity (Figure 5E,F). The same patterns held when splitting the participants based on the other functional measures (Supplementary table 2).

**Figure 5:**
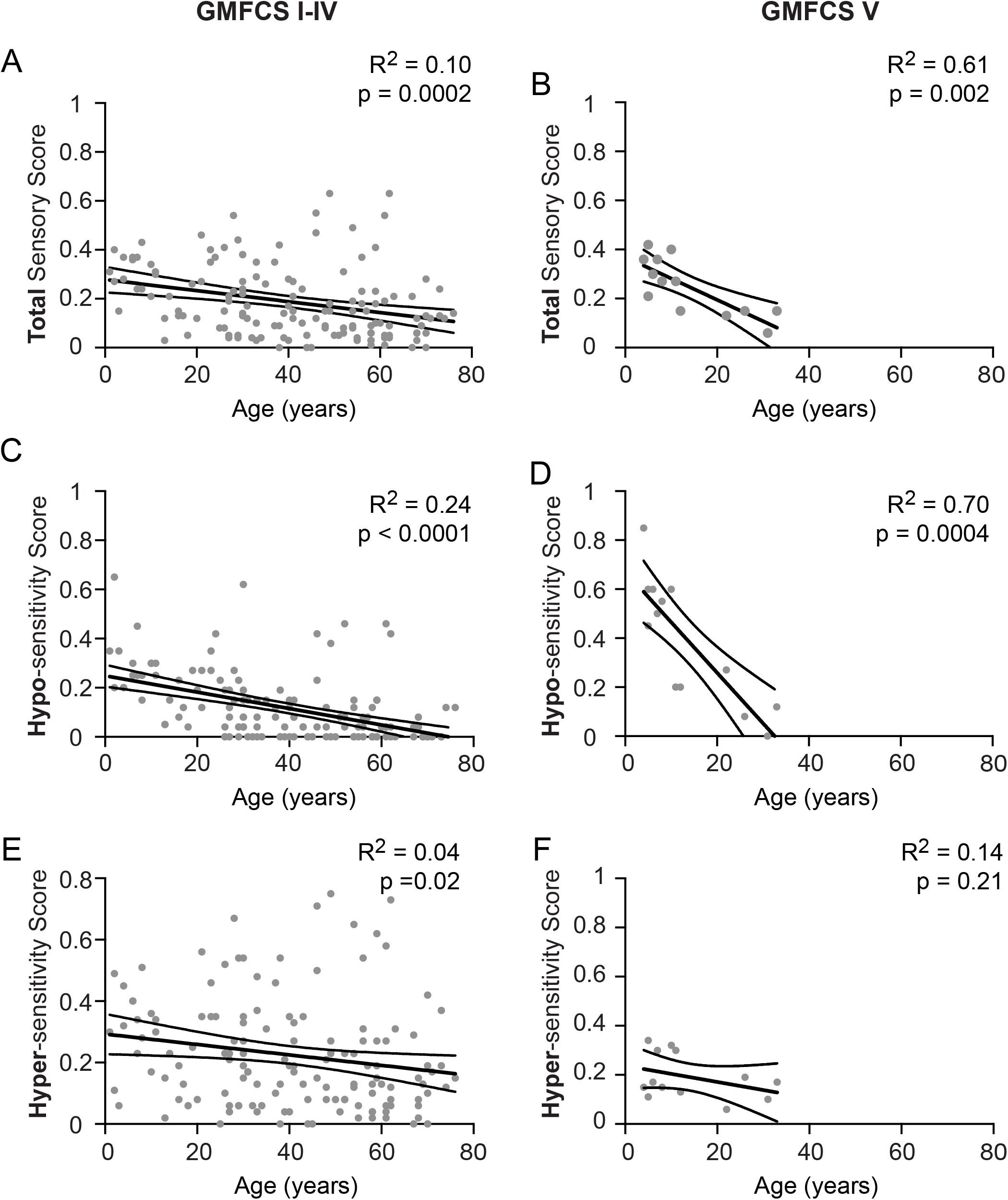
Sensory symptoms decrease with age for both high and low GMFCS groups. Total sensory (A, B), Hypo-sensitivity (C,D) and Hyper-sensitivity (E,F) scores decreased with age for individuals with GMFCS I-IV as well as individuals with GMFCS V. Of note, the association between sensory symptoms and age are shown separately for GMFCS Levels I-IV and GMFCS Level V because the only significant difference in age between GMFCS levels was between Levels I-IV and Level V (see Figure 1). Pearson correlation, *p<0.05.

Having shown that age dependent decreases in hyposensitivity were most prominent for individuals with the greatest functional limitations, we wondered how sensory modality specific changes correlated to related functional areas. For example, prior work suggested aberrant tactile perception drives mobility impairments^26^. Interestingly, tactile hypo-sensitivity had the strongest correlation with age (R^2^ = 0.29, Figure 6A) as compared to hyposensitivity related to the vision (R^2^= 0.17, Figure 6B), auditory (R^2^= 0.11, Figure 6C) and proprioception (R^2^= 0.11, Figure 6D) modalities.

**Figure 6:**
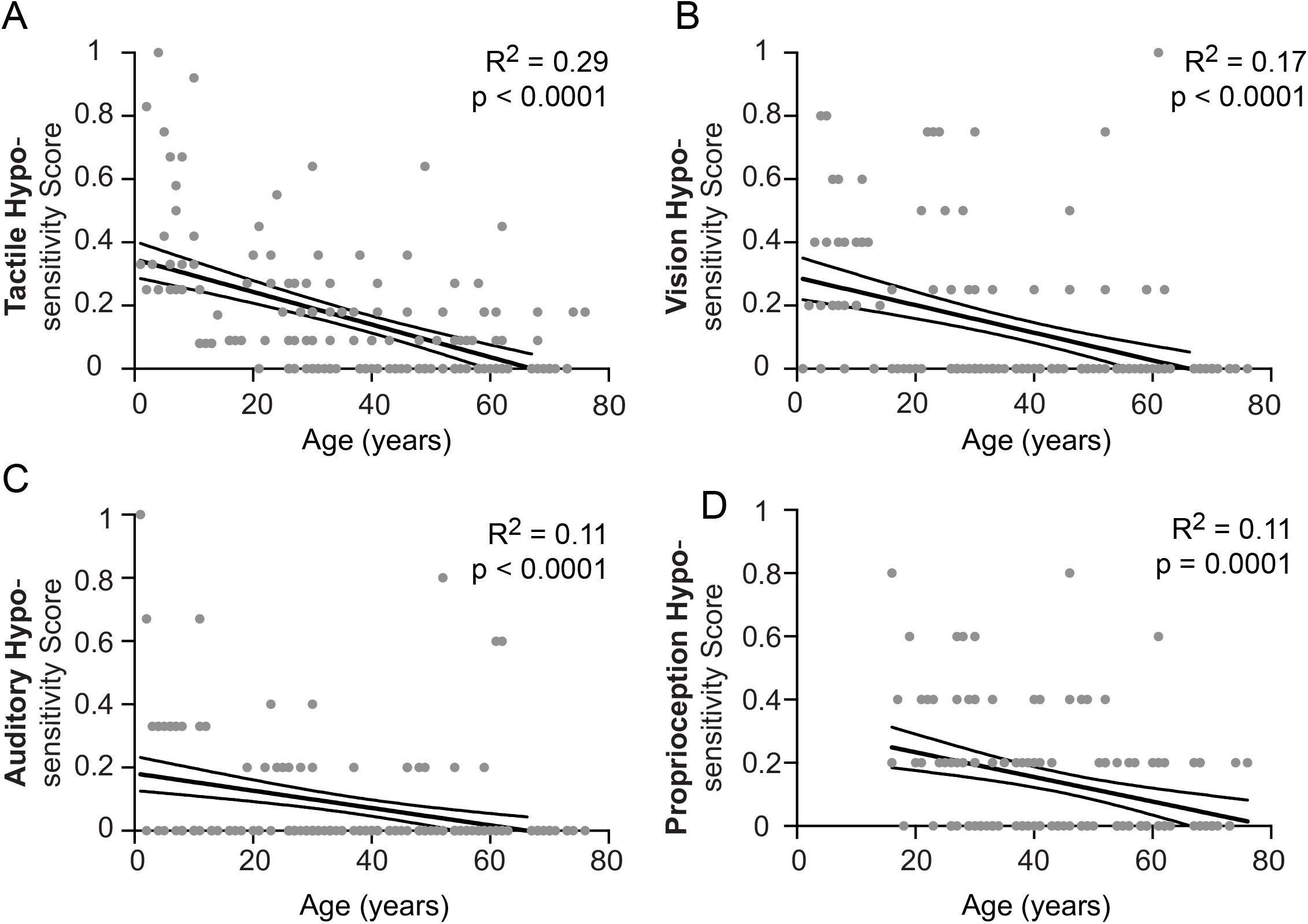
Sensory symptoms decrease with age across all modalities. Tactile (A), Vision (B), Auditory (C) and Proprioceptive (D) hypo-sensory scores decreased with age. Pearson correlation, *p<0.05.

Given that sensory symptoms, functional level, and age all appeared to be correlated with each other in the respondent pool, we also assessed whether age and functional classification system levels could predict SPS-I scores (total, hypo-sensitivity, and hyper-sensitivity) using multiple linear regression analysis. Multiple linear regression models including all 6 variables (age plus five classification system levels) were able to significantly predict total SPS-I score (F=5.9, p<0.0001), hyposensitivity SPS-I score (F=24.8, p<0.0001) and the hypersensitivity SPS-I score (F=2.2, p<0.0001) in our dataset. VIFs were all less than 5 (range 1.3-2.1) suggesting a lack of multi-collinearity between the predictive variables. Of note, taking all 6 variables into account, age was a significant predictor of total SPS-I score (p=0.003) and the hypo-sensitivity SPS-I score (p<0.0001), but not the hyper-sensitivity SPS-I score (p=0.08). This further suggests that decreased hypo-sensitivity symptoms, more so than decreased hyper-sensitivity symptoms, drives decreased SPS-I score seen with increased age.

## Discussion

The primary finding of this work is that sensory symptoms, particularly tactile hypo-sensitivity, decrease with age in individuals with CP. Interestingly, age dependent changes in sensory symptoms have been reported in Autism with mixed findings. In a study of 55 children with Autism and 35 children with other neurodevelopment disabilities, hypo-responsiveness declined in children with Autism but not other developmental disabilities while both groups exhibited an age dependent decline in sensory seeking behaviors^27^. In contrast, no decreases in sensory symptoms were seen in either 29 children with Autism or 26 children with other neurodevelopmental disabilities followed from age 2 to 8 years^28^. In the largest study to date, model-based cluster analysis of sensory symptoms from 919 children with Autism age 3 to 14 suggested that only hyper-sensitivity was associated with age^29^. Thus, decreases in hyposensitivity with age are likely specific to cerebral palsy and merit further investigation.

### Implications of age-related sensory changes in people with CP

We show that sensory symptoms in people with CP ameliorate with age, yet gross motor function declines with age, with peak gross motor function in early adulthood^13^. This raises the question: how do sensory symptoms relate to functional deficits in CP? We have shown that people with greater functional impairments across all assessed modalities have a greater number of sensory symptoms. Though these relationships are correlational, it is likely that altered sensation deleteriously affects function. For example, somatosensory cortical lesions are associated with upper extremity functional limitation, proposed to be independent from and additive to pure motor impairments^30^. Sensory symptoms similarly correlate with functional ability in autism^31^.

Tactile hyposensitivity – the most commonly affected modality in our study – is directly related to function. For example, the brain relies on tactile feedback during grasping and object manipulation to adapt fingertip forces to the object being moved^32^ and thus tactile hyposensitivity could lead to the decreased precision grip in CP. Similarly, spatial tactile deficits account for ~30 percent of variance in upper limb motor function in children with unilateral CP^33^. Thus, there is increasing evidence for a causative role of sensory symptoms in motor function for which greater understanding of the patterns of sensory symptoms in adults with CP will be important.

Other functionally limiting symptoms, like pain, also worsen with age in patients with CP^14^. While multiple factors contribute to worsening pain with age, our findings raise an interesting possibility that the high rates of hyposensitivity observed in young people may be protective against symptoms like pain that emerge once sensory symptoms resolve. Sensory symptoms could also directly affect pain perception at the level of the somatosensory, insular, and cingulate cortices^34–35^. Investigating the mechanistic links between age-related hyposensitivity decreases and increased pain may inform pain therapeutics targeted specifically for people with CP.

### A role for tactile based therapy in people with CP

Given the proposed connections between sensory symptoms and functional ability, sensory-focused therapeutics are being proposed to improve sensorimotor integration and movement function^36^. The correlations in the tactile modality between hypo-sensitivity and age found in this study support a prospective approach of occupational therapy, massage, and tactile sensory stimulation to improve motor function in individuals with CP.

Sensory integration therapy is often incorporated into traditional occupational therapy, a mainstay in the management of CP. For example, dynamic surface exercise therapy allows children to get proprioceptive and sensory input while completing motor tasks. It is theorized that minute, but frequent postural perturbations of the dynamic surface strengthen trunk musculature, improving balance and righting maneuvers more effectively than performing the same exercises on a static surface^37^. Foot and ankle sensory symptoms may also have an effect on balance and gross motor function which may be improved with physical therapy techniques like stochastic resonance stimulation.^38^ To fully understand the role of therapies targeting sensory symptoms, further work characterizing sensory phenotypes in people with CP – particularly adults - is needed.

### Limitations

This study has several limitations. First, any measure of a subjective behavior, including the well-validated SPS-I, has limitations. The SPS-I is a self or caregiver report measure which is a common but indirect assessment of sensory function. Therefore, quantitative sensory testing of adults with CP will be important to confirm these findings. The granularity of comparisons within and between sensory modalities using the SPS-I may also be affected by the number of questions used to assess each modality, though we did correct within-modality scores based on the number of questions used to assess that modality. Furthermore, the novel aggregate analysis of the SPS-I sub-scores across both pediatric and adult age groups as done for this study would benefit from independent validation in a separate population. Secondly, our survey, as is true for many virtual surveys, skews toward females,^39^ even though there are more males than females with CP at large.^40^ Thirdly, as we collected data through the MyCP network, we do not have a control population and “typical” cut-off values are only available for pediatric populations. Therefore, it is not possible to separate sensory symptoms related to aging that are independent of CP. Finally, our population was skewed with higher GMFCS in the younger age group and may have been skewed toward people with CP who chose to respond because they experience sensory symptoms. While this is the largest sensory study in people with CP to date, future studies should aim for even larger groups with data collected in people across all functional status in a comprehensive and prospective manner.

## Conclusion

In conclusion, by leveraging the participation of the CPRN, we characterize patterns of sensory changes in individuals with CP and find a wide range of sensory differences across ages and functional levels impacting all sensory modalities. Further, we show that older people with CP have fewer sensory symptoms than younger people with CP and that this effect is most pronounced in people with the greatest functional impairments. Notably, most prior work evaluating sensory symptoms in CP has been in children, but this work demonstrates the critical importance of including adults with CP in future sensory studies. These findings are important when considering treatments for people with CP for whom sensory symptoms should be systematically assessed.

## Data Availability

All data produced in the present study are available upon reasonable request to the authors

## Abbreviations

CP: cerebral palsy
REDCap: research electronic data capture
SPS-I: Sensory Processing Scale Inventory
ANOV: analysis of variance
IRB: institutional review board
CPRN: Cerebral Palsy Research Network
GMFCS: Gross Motor Function Classification System
MACS: Manual Ability Classification System)
EDACS: Eating and Drinking Ability Classification System
CFCS: Communication Function Classification System
VFCS: Visual Function Classification System

## Acknowledgements

We acknowledge and thank Dr. Sarah Schoen for allowing the use of the Sensory Processing Scale Inventory. We also thank MyCP Research Network for facilitating data collection.

---

**Supplementary Table 1:**
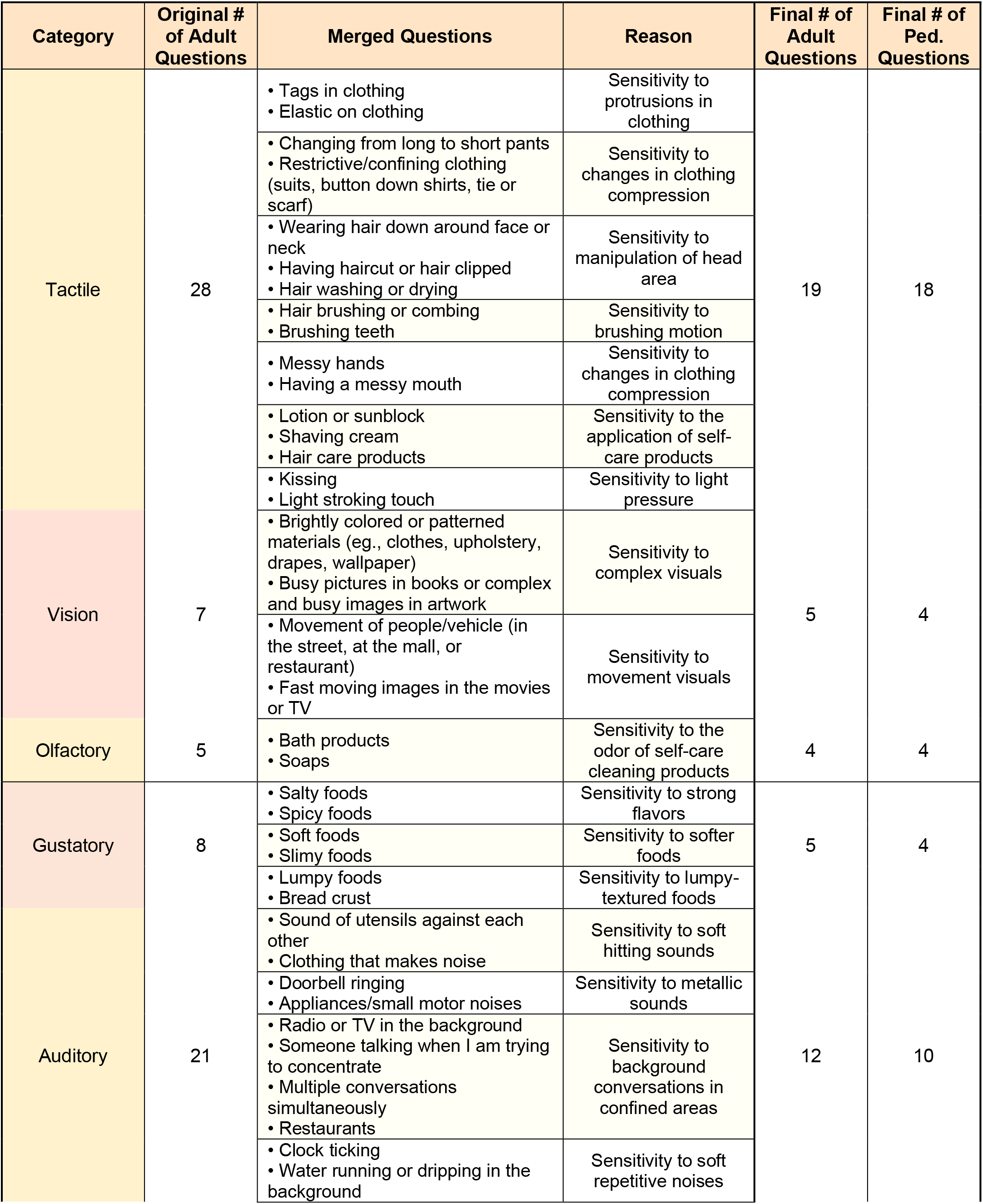
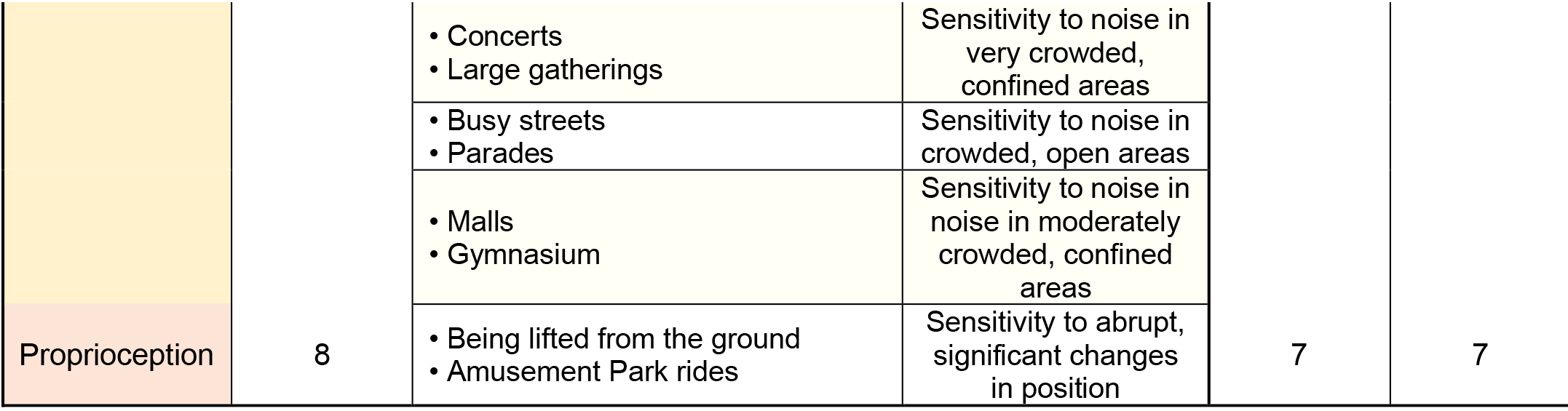
Modified analysis of the Sensory Processing Scale Inventory For each sensory modality, qualitatively similar questions from the adult survey were merged to allow for similar numbers of adult and pediatric questions. The specific questions that were merged are shown.

**Supplementary Table 2:** Correlation between sensory score and age based on functional group Sensory symptoms, particularly Hypo-sensitivity, decrease with age in individuals in both high and low functional classification systems. For each of these four functional classification systems, groups were divided into functional level I (least impairment) vs. levels II-V (more impairment) because hyposensitivity scores were lower in individuals in functional level I (see Figure 4). In both functional categories and across all functional scales, hypo-sensitivity score improved with age. Multiple linear regression, *p<0.05. MACS = Manual Ability Classification System, CFCS = Communication Function Classification System, EDACS = Eating and Drinking Ability Classification System, VFCS = Visual Function Classification System.

|  |  | Total Sensory Score<br>vs. Age |  | Hypo-sensitivity Score<br>vs. Age |  | Hyper-sensitivity Score<br>vs. Age |  |
| --- | --- | --- | --- | --- | --- | --- | --- |
|  |  | R<sup>2</sup> | p-value | R<sup>2</sup> | p-value | R<sup>2</sup> | p-value |
| **MACS** | MACS I | 0.07 | 0.01 | 0.18 | <0.0001 | 0.03 | 0.10 |
|  | MACS II-V | 0.11 | 0.004 | 0.31 | <0.0001 | 0.02 | 0.30 |
| **CFCS** | CFCS I | 0.10 | 0.0008 | 0.25 | <0.0001 | 0.04 | 0.03 |
|  | CFCS II-V | 0.03 | 0.26 | 0.19 | 0.003 | <0.01 | 0.81 |
| **EDACS** | EDACS I | 0.10 | 0.001 | 0.27 | <0.0001 | 0.03 | 0.07 |
|  | EDACS II-V | 0.03 | 0.23 | 0.25 | 0.0003 | <0.01 | 0.78 |
| **VFCS** | VFCS I | 0.12 | <0.0001 | 0.28 | <0.0001 | 0.04 | 0.02 |
|  | VFCS II-V | <0.01 | 0.93 | 0.13 | 0.04 | 0.02 | 0.45 |

